# Longitudinal evaluation and decline of antibody responses in SARS-CoV-2 infection

**DOI:** 10.1101/2020.07.09.20148429

**Authors:** Jeffrey Seow, Carl Graham, Blair Merrick, Sam Acors, Kathyrn J.A. Steel, Oliver Hemmings, Aoife O’Bryne, Neophytos Kouphou, Suzanne Pickering, Rui Pedro Galao, Gilberto Betancor, Harry D. Wilson, Adrian W. Signell, Helena Winstone, Claire Kerridge, Nigel Temperton, Luke Snell, Karen Bisnauthsing, Amelia Moore, Adrian Green, Lauren Martinez, Brielle Stokes, Johanna Honey, Alba Izquierdo-Barras, Gill Arbane, Amita Patel, Lorcan O’Connell, Geraldine O’Hara, Eithne MacMahon, Sam Douthwaite, Gaia Nebbia, Rahul Batra, Rocio Martinez-Nunez, Jonathan D. Edgeworth, Stuart J.D. Neil, Michael H. Malim, Katie J Doores

## Abstract

Antibody (Ab) responses to SARS-CoV-2 can be detected in most infected individuals 10-15 days following the onset of COVID-19 symptoms. However, due to the recent emergence of this virus in the human population it is not yet known how long these Ab responses will be maintained or whether they will provide protection from re-infection. Using sequential serum samples collected up to 94 days post onset of symptoms (POS) from 65 RT-qPCR confirmed SARS-CoV-2-infected individuals, we show seroconversion in >95% of cases and neutralizing antibody (nAb) responses when sampled beyond 8 days POS. We demonstrate that the magnitude of the nAb response is dependent upon the disease severity, but this does not affect the kinetics of the nAb response. Declining nAb titres were observed during the follow up period. Whilst some individuals with high peak ID_50_ (>10,000) maintained titres >1,000 at >60 days POS, some with lower peak ID_50_ had titres approaching baseline within the follow up period. A similar decline in nAb titres was also observed in a cohort of seropositive healthcare workers from Guy’s and St Thomas’ Hospitals. We suggest that this transient nAb response is a feature shared by both a SARS-CoV-2 infection that causes low disease severity and the circulating seasonal coronaviruses that are associated with common colds. This study has important implications when considering widespread serological testing, Ab protection against re-infection with SARS-CoV-2 and the durability of vaccine protection.

## Introduction

Severe acute respiratory syndrome coronavirus 2 (SARS-CoV-2) is a betacoronavirus responsible for coronavirus disease-19 (COVID-19). Spike (S) is the virally encoded surface glycoprotein facilitating angiotensin converting enzyme-2 (ACE-2) receptor binding on target cells through its receptor binding domain (RBD). In a rapidly evolving field, researchers have already shown that, in most cases, individuals with a confirmed PCR diagnosis of SARS-CoV-2 infection develop IgM, IgA and IgG against the virally encoded surface spike protein (S) and nucleocapsid protein (N) within 1-2 weeks post onset of symptoms (POS) and remain elevated following initial viral clearance.^1-7^ S is the target for nAbs, and a number of highly potent monoclonal antibodies (mAbs) have been isolated that predominantly target the RBD.^8,9^ A wide range of SARS-CoV-2 neutralizing antibody (nAb) titres have been reported following infection and these vary depending on the length of time from infection and the severity of disease.^4^ Further knowledge on the magnitude, timing and longevity of nAb responses following SARS-CoV-2 infection is vital for understanding the role nAbs might play in disease clearance and protection from re-infection (also called renewed or second wave infections). Further, as a huge emphasis has been placed on serological assays to determine seroprevalence against SARS-CoV-2 in the community and estimating infection rates, it is important to understand immune responses following infection to define parameters in which Ab tests can provide meaningful data in the absence of PCR testing in population studies.

Ab responses to other human coronaviruses have been reported to wane over time.^10-13^In particular, Ab responses targeting endemic human alpha- and betacoronaviruses can last for as little as 12 weeks,^14^ whereas Abs to SARS-CoV and MERS can be detected in some individuals 12-34 months after infection.^11,15^ Although several cross-sectional studies of nAb responses arising from SARS-CoV-2 infection have been reported,^4,7^ there is currently a paucity of information on the longevity of the nAb response using multiple sequential samples from individuals in the convalescent phase beyond 30-40 days POS.^3,5,16^ This study uses sequential samples from 65 individuals with PCR confirmed SARS-CoV-2 infection and 31 seropositive healthcare workers (HCW) up to 94 days POS to understand the kinetics of nAb development and the magnitude and durability of the nAb response.

Here, we measured the Ab binding response to S, the receptor binding domain (RBD) and N, as well as the neutralization potency against SARS-CoV-2 using an HIV-1 based pseudotype assay. We show that IgM and IgA binding responses decline after 20-30 days POS. We demonstrate that the magnitude of the nAb response is dependent upon the disease severity but this does not impact on the time to ID_50_ peak (serum dilution that inhibits 50% infection). nAb titres peak on average at day 23 POS and then decrease 2- to 23-fold during an 18-65 day follow up period. In individuals that only develop modest nAb titres following infection (100-300 range), titres become undetectable (ID_50_ <50) or are approaching baseline after ∼50 days highlighting the transient nature of the Ab response towards SARS-CoV-2 in some individuals. In contrast, those with high peak ID_50_ for neutralization maintain nAb titres in the 1000-3500 range at the final timepoint tested (>60 days POS). This study has important implications when considering protection against re-infection with SARS-CoV-2 and the durability of vaccine protection.

## Results

### Cohort description

The antibody response in 65 RT-qPCR confirmed SARS-CoV-2-infected individuals was studied over sequential time points. The cohort consisted of 59 individuals admitted to, and 6 healthcare workers (HCW) at, Guy’s and St Thomas’ NHS Foundation Trust (GSTFT). The cohort were 77.2% male with average age of 55.2 years (range 23-95 years). Ethnicity information was not collected on this cohort. A severity score was assigned to patients based on the maximal level of respiratory support they required during their period of hospitalisation. The score, ranging from 0-5 (see methods), was devised to mitigate underestimating disease severity in patients not for escalation above level one (ward-based) care. This cohort included the full breadth of COVID-19 severity, from asymptomatic infection to those requiring extra corporeal membrane oxygenation (ECMO) for severe respiratory failure. Comorbidities included diabetes mellitus, hypertension, and obesity, with a full summary in **Table S1**. Sequential serum samples were collected from individuals at time-points between 1-and 94-days post onset of symptoms (POS) and were based upon availability of discarded samples taken as part of routine clinical care, or as part of a HCW study.

### Antibody binding responses to SARS-CoV-2

The IgG, IgM and IgA response against spike (S), the receptor binding domain (RBD) and nucleocapsid (N) were measured by ELISA over multiple time points (**Figure 1 and S1**).^6^ Initially, the optical density at 1:50 serum dilution was measured for 300 samples from the 65 individuals (**Figure 1 and S1**). Only 2/65 individuals (3.1%) did not generate a detectable Ab response against any of the antigens in the follow up period (**Table S2**). However, sera were only available up until 2- and 8-days POS for these two individuals and as the mean time to seroconversion against at least 1 antigen was 12.6 days POS, it is likely these individuals may have seroconverted at a later time point after they were discharged from hospital. IgG responses against S, RBD and N antigens were observed in 92.3%, 89.2% and 93.8% of individuals respectively (**Table S2**). The frequency of individuals generating an IgM response was similar to IgG, with 92.3%, 92.3% and 95.4% seropositive against S, RBD and N respectively. The frequency of individuals with an IgA response to RBD and N was lower, with only 72.3% and 84.6% seropositive respectively (**Table S2**) whereas the IgA to S frequency was similar to the IgM and IgG.

**Figure 1:**
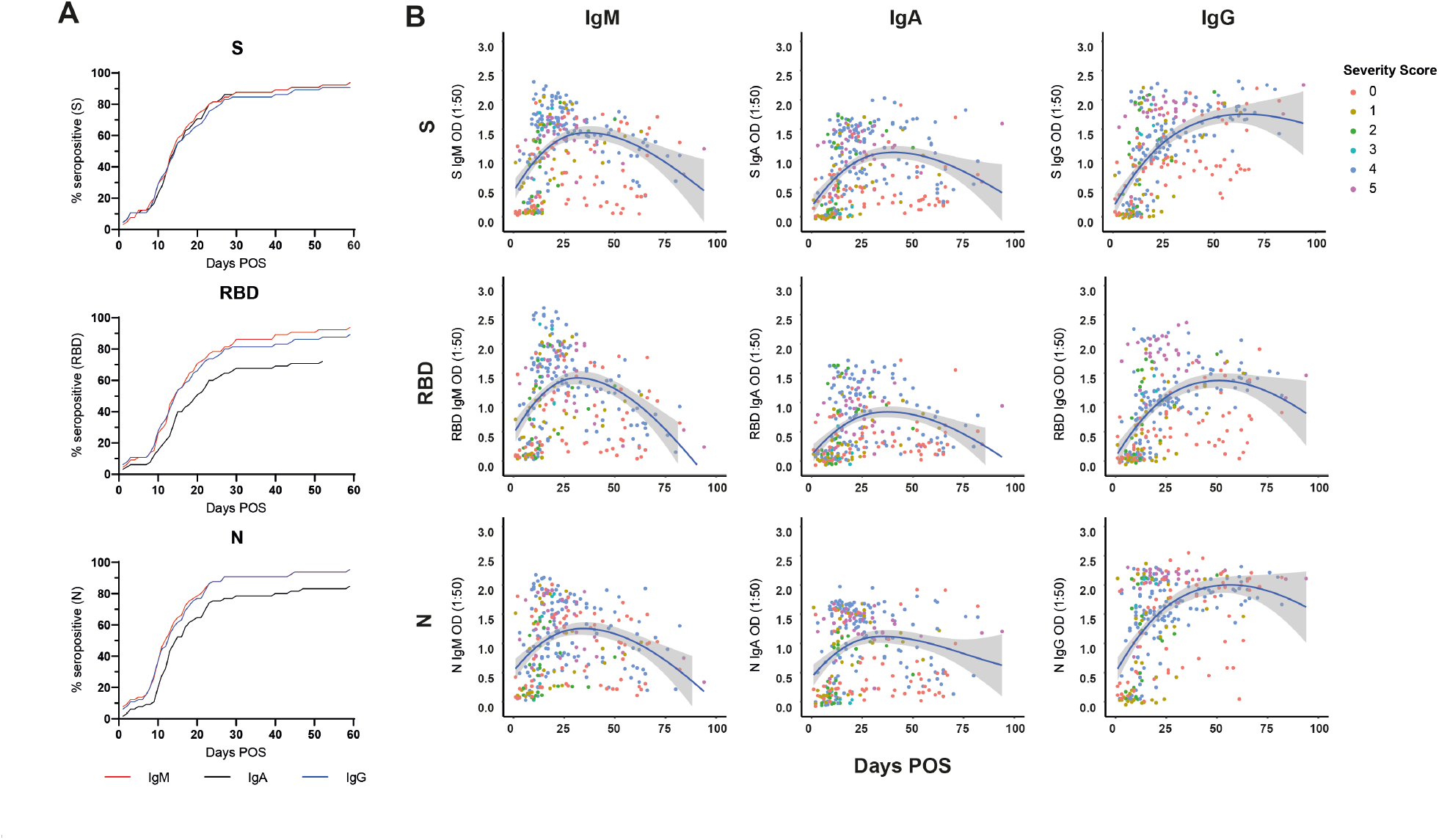
Kinetics of antibody development against SARS-CoV-2 antigens over time. A) A cumulative frequency analysis describing the point of seroconversion for each person in the cohort. Graph shows the percentage of individuals in the cohort that become IgM, IgA or IgG positive to S, RBD and N each day. A serum is considered positive when the OD is 4-fold above background. B) IgM, IgA and IgG OD values against S, RBD and N are plotted against the time post onset of symptoms (POS) at which sera was collected. Coloured dots indicate disease severity (0-5). The line shows the mean OD value expected from a Loess regression model, the ribbon indicates the pointwise 95% confidence interval. OD = optical density.

A cumulative frequency analysis of positive IgG, IgA and IgM responses against S, RBD and N across the cohort did not indicate a more rapid elicitation of IgM and IgA responses against a particular antigen (**Figure 1A and S2A**) and may reflect the sporadic nature in which sequential serum samples were collected. Therefore, a subset of donors from whom sera was collected over sequential time points early in infection (<14 days POS) were analysed further and different patterns of seroconversion were observed (**Figure S2B**). 51.6% (16/31) of individuals showed synchronous seroconversion to IgG, IgM and IgA whilst some individuals showed singular seroconversion to IgG (9.7%), IgM (9.7%) and IgA (9.7%). 58.1% (18/31) of individuals showed synchronous seroconversion to S, RBD and N, whereas singular seroconversion to N or S were both seen in 16.1% of individuals.

Longitudinal analysis across sequential samples highlighted the rapid decline in the IgM and IgA response to all three antigens following the peak OD between 20- and 30-days POS for IgM and IgA respectively (**Figure 1B and S1A**) as might be expected following an acute infection. For some individuals sampled at time points >60 days POS, the IgM and IgA responses were approaching baseline (**Figure 2B and S1A**). In contrast, the IgG OD (as measured at 1:50 dilution) remained high in the majority of individuals, even up to 94 days POS (**Figure 1B and S1A**). However, differences were apparent when patients were stratified by disease severity and when half maximal binding (EC_50_) was measured (see below).

**Figure 2:**
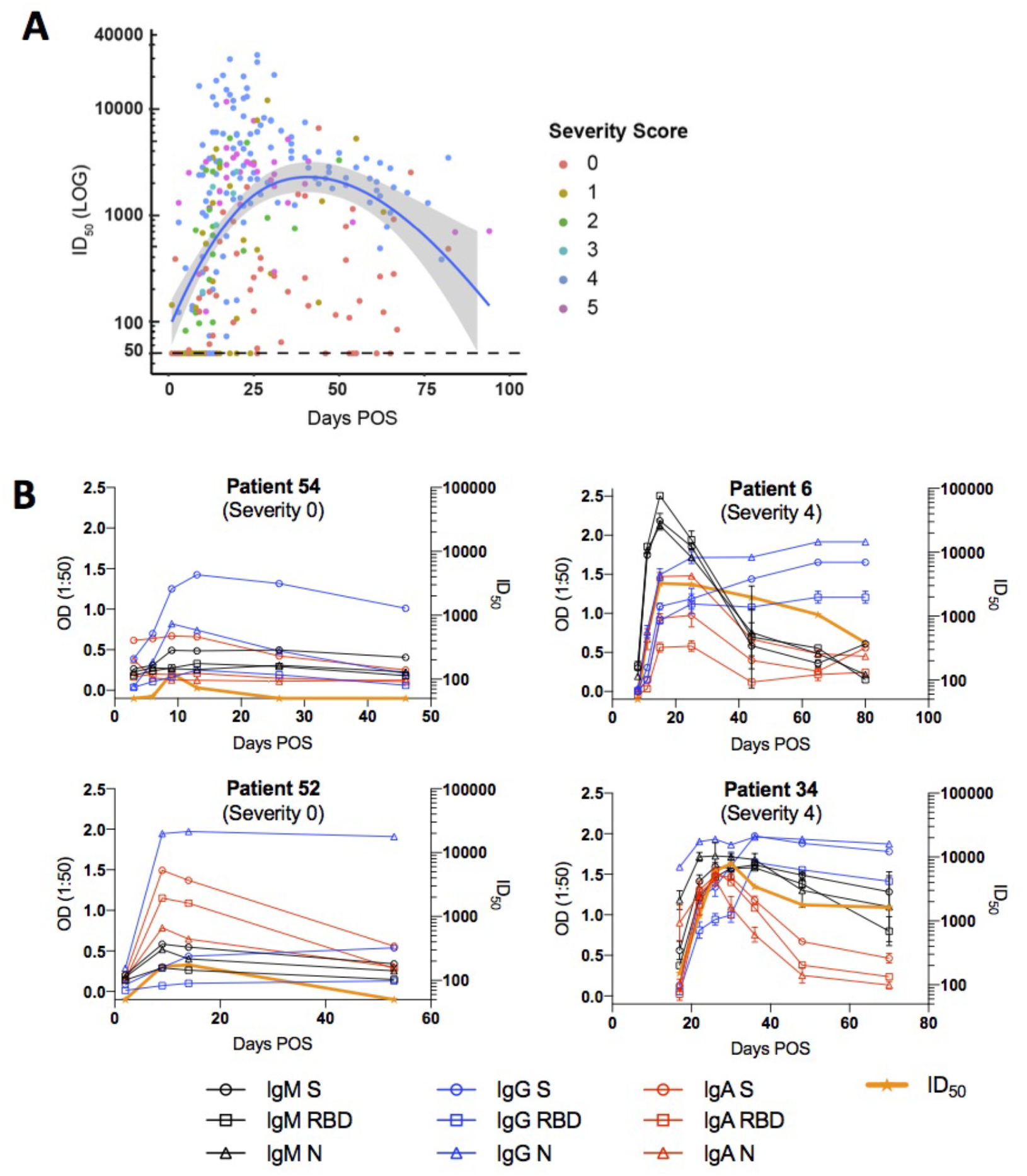
Kinetics of neutralizing antibody responses in SARS-CoV-2 infection. A) ID_50_ values plotted against the days post onset of symptoms (POS) at which sera was collected. Coloured dots indicate disease severity (0-5). The line shows the mean ID_50_ value expected from a Loess regression model, the ribbon indicates the pointwise 95% confidence interval. B) Example kinetics of Ab responses for four individuals during acute infection and the convalescent phase. Graphs show comparison between severity 0 (left) and severity 4 (right) rated disease.

### Neutralizing antibody responses to SARS-CoV-2

We next measured SARS-CoV-2 neutralization potency using HIV-1 (human immunodeficiency virus-1) based virus particles, pseudotyped with SARS-CoV-2 S^17,18^ in a HeLa cell line stably expressing the ACE2 receptor. Increased neutralization potency was observed with increasing days POS (**Figure 2A**) with each individual reaching a peak neutralization titre (ranging from 98 to 32,000) after an average of 23.1 days POS (range 1-66 days) (**Figure S1B**). Only two individuals (3.1%) did not develop a nAb response (ID_50_ <50) which was consistent with their lack of binding Abs at the time points tested (<8 days POS). At peak neutralization, 7.7% had low (50-200), 10.8% medium (201-500), 18.5% high (501-2000) and 60.0% potent (2001+) neutralizing titres. For serum samples collected after 65 days POS, the percentage of donors with potent nAbs (ID_50_>2000) had reduced to 16.7% (**Table S3**). Neutralization ID_50_ values correlated well with IgG, IgM and IgA binding OD values to all three antigens, S, RBD and N (**Figure S3**), and the best fit (r^2^) was observed between ID_50_ and the OD for S IgA and S IgM. The average time to detectable neutralization was 14.3 days POS (range 3-59 days). At earlier time points POS, some individuals displayed neutralizing activity before an IgG response to S and RBD was detectable by ELISA (**Figure S2C**). This highlights the capacity of S- and RBD-specific IgM and IgA in acute infection to facilitate neutralization in the absence of measurable IgG.^19^

To determine how disease severity impacts Ab titres, we compared the ID_50_ values between individuals with 0-3 disease severity with those in the 4/5 group (**Figure 3**). Although the magnitude of the nAb response at peak neutralization was significantly higher in the severity 4/5 group (**Figure 3A**), the time taken to measure detectable nAb titres (**Figure 3C**) and the time of peak neutralization (**Figure 3B**) did not differ between the two groups suggesting disease severity enhances the magnitude of the Ab response but does not alter the kinetics. Comparison of the IgG, IgM and IgA OD values against S at peak neutralization showed significantly higher IgA and IgM ODs in the severity 4/5 group but no significant difference was observed for IgG to S (**Figure 3D-F**). This observation may further highlight a potential role for IgA and IgM in neutralization.^19^ Within the severity 4/5 group, a proportion of patients were treated with immunomodulation for a persistent hyperinflammatory state characterized by fevers, markedly elevated CRP and ferritin, and multi-organ dysfunction. Despite an initial working hypothesis that antibody responses may differ either as a cause or consequence of this phenotype, no difference in ID_50_ titres was observed between these individuals and the remainder of the severity 4/5 cases (**Figure 3G**).

**Figure 3:**
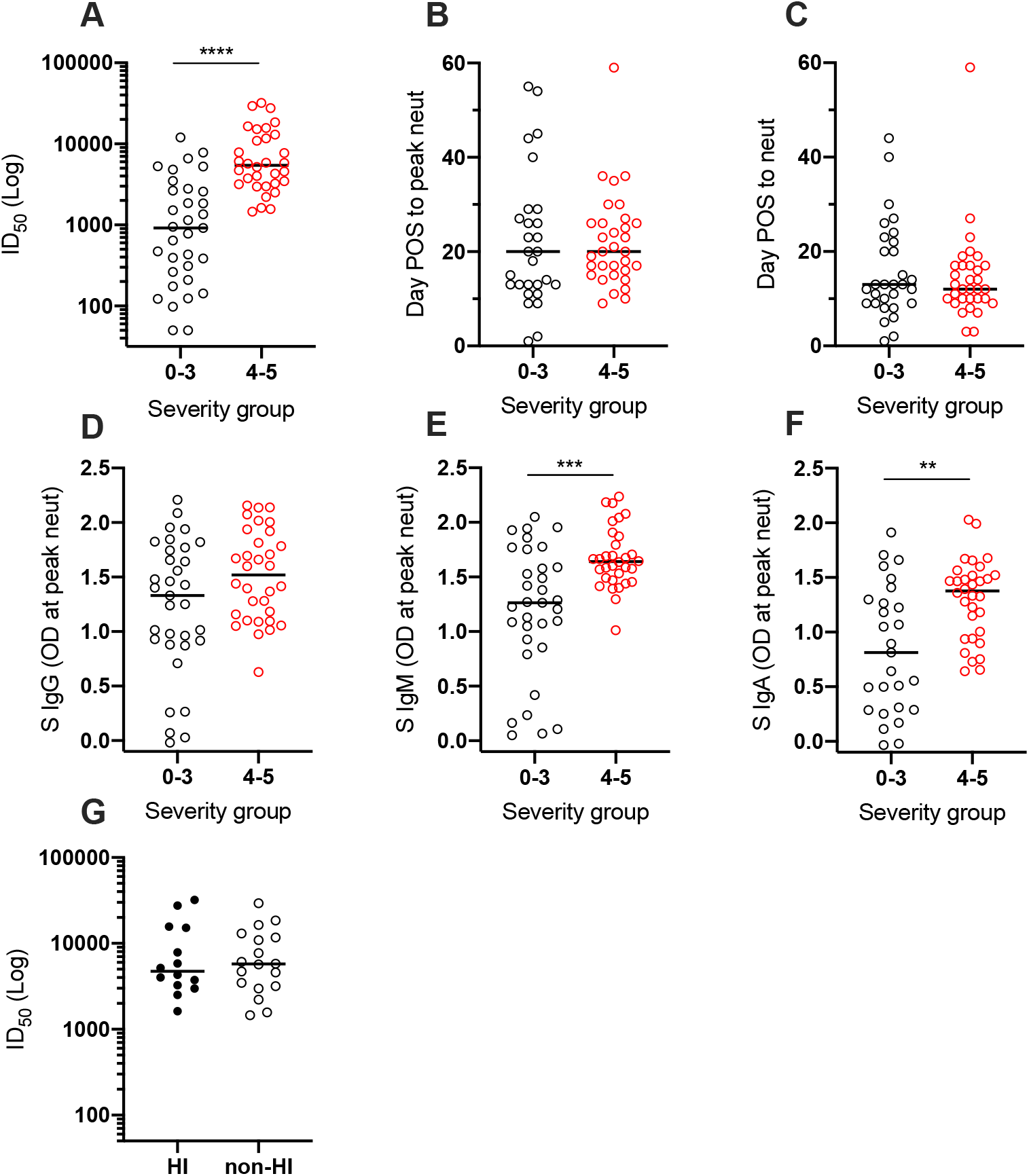
Impact of disease severity of Ab responses to SARS-CoV-2 infection. Comparison for individuals with 0-3 or 4/5 disease severity for A) peak ID_50_ of neutralization (*p*<0.0001), B) the time POS to reach peak ID_50_ (*p*=0.674), and C) the time POS to detect neutralizing activity (*p*=0.9156). Comparison in OD values for individuals with 0-3 or 4/5 disease severity for D) IgG (*p*=0.0635), E) IgM (*p*=0.0003) and F) IgA (*p*=0.0018) against S measured at peak ID_50_. G) Comparison of the peak ID_50_ value for individuals who were treated for hyperinflammation or not, and had 4/5 disease severity (*p*>0.999). Statistical significance was measured using a Mann-Whitney test.

### Longevity of the Ab response

Following the peak in neutralization, a waning in ID_50_ was detected in individuals sampled at >40 days POS. Comparison of the ID_50_ at peak neutralization and ID_50_ at the final time point collected showed a decrease in almost all cases (**Figure 4A**). For some individuals with severity score 0, where the peak in neutralization was in the ID_50_ range 100-300, neutralization titres became undetectable (ID_50_ <50) in the pseudotype neutralization assay at subsequent time points (**Figure 4A** and **2B**). For example, donors 52 and 54 both generated a low nAb response (peak ID_50_ of 174 and 434 respectively) but no neutralization could be detected in our assay 39 and 34 days after the peak in ID_50_ respectively (**Figure 2B**).

**Figure 4:**
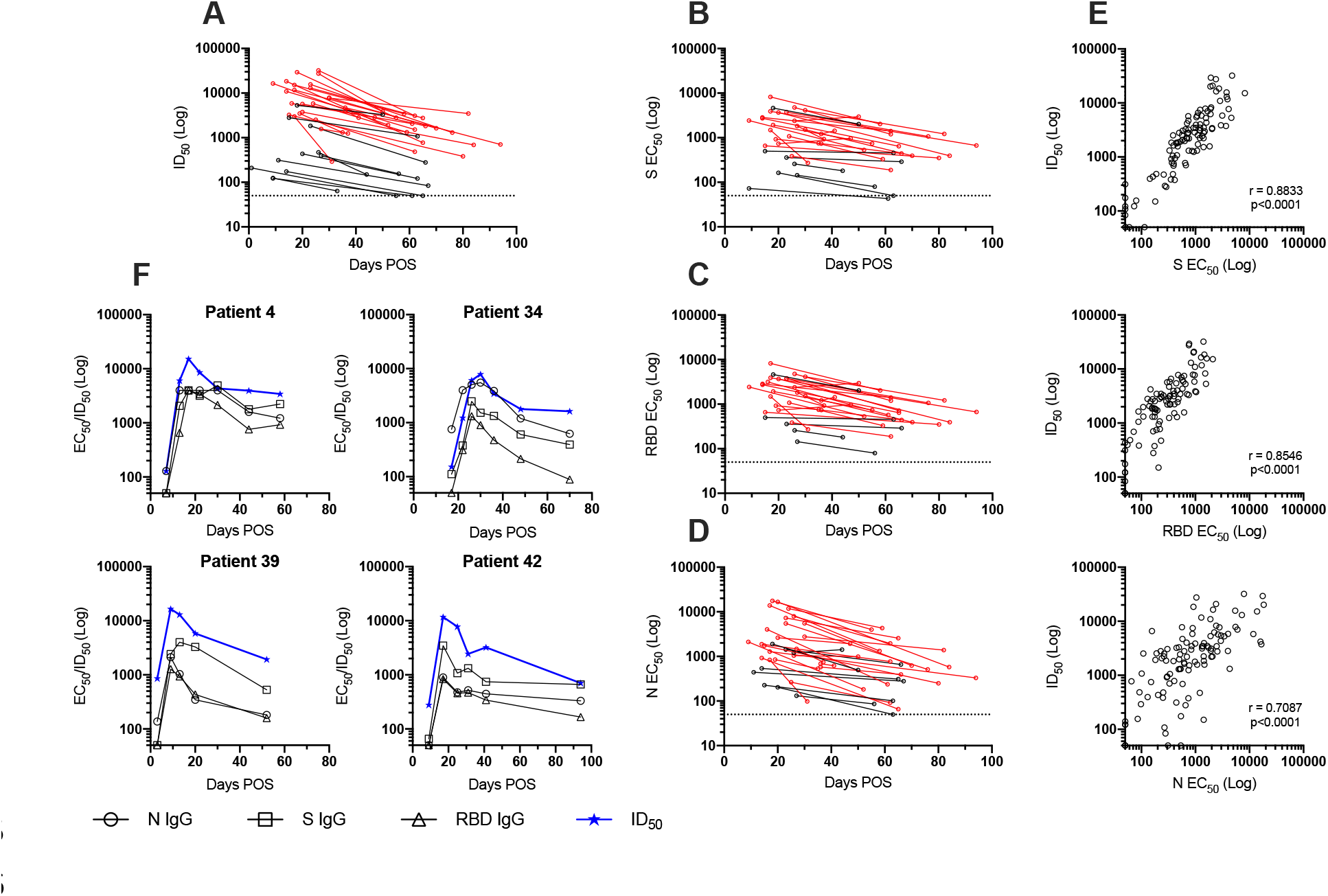
Longevity of the Ab response. A) ID_50_ at peak neutralization is plotted against the donor matched ID_50_ at the last time point sera was collected. Only individuals where the peak ID_50_ occurs before the last time point, and where the last time point is >30 days POS are included in this analysis. B-D) EC_50_ values for IgG binding to S, RBD and N were calculated at time point with peak ID_50_ and the final time point. EC_50_ at peak neutralization is plotted with the donor matched EC_50_ at the last time point sera was collected. Individuals with a disease severity 0-3 are shown in black and those with 4/5 are shown in red. E) Correlation of ID_50_ with IgG EC_50_ against S (*r*^*2*^=0.8293), RBD (*r*^*2*^=0.7128) and N (*r*^*2*^=0.4856) (Spearman correlation, *r*. A linear regression was used to calculate the goodness of fit, *r*^*2*^). F) Change in IgG EC_50_ measured against S, RBD and N and ID_50_ over time for 4 example patients (all severity 4).

To gain a more quantitative assessment of the longevity of the IgG binding titres specific for S, RBD and N, EC_50_ values were measured at the peak of neutralization and compared to the EC_50_ at the final time point collected. EC_50_ values correlated very well with ID_50_ (**Figure 4E**). Similar to neutralization potency, a decrease in EC_50_ was observed within the follow up period for S, RBD and N (**Figure 4B-D**). For those whose nAb titre decreased towards baseline, the EC_50_ for IgG to S and RBD also decreased in a similar manner. Finally, to determine whether the reduction in IgG titres might plateau, EC_50_ values for all time points for four representative individuals were measured who had multiple samples collected in the convalescent phase (**Figure 4F**). A steady decline in neutralization was accompanied by a decline in IgG binding to all antigens within the time window studied. Further assessment of Ab binding and neutralizing titres in samples collected >94 days POS will be essential to fully determine the longevity of the nAb response.

### Ab responses in a Healthcare worker cohort

To gain further understanding of Ab responses in SARS-CoV-2 infection we next analysed sequential serum samples from 31 seropositive (as determined by an IgG response to both N and S)^6^ healthcare workers (HCW) from GSTFT. Ab responses in these individuals are likely to be more akin to those who were never hospitalised. Sera were collected every 1-2 weeks from March - June 2020 and any symptoms relating to COVID-19 recorded. Acute infection, as determined by detectable SARS-CoV-2 RNA on RT-qPCR, was not measured routinely. 80.6% (25/31) of seropositive individuals recorded COVID-19 compatible symptoms (including fever, cough and anosmia) since 1^st^ February 2020, 19.4% (6/31) reported none.

IgG and IgM binding to S, RBD and N by ELISA and neutralization titres were measured over time using sequential samples (**Figure 5A and S4A**). Similar to the patient cohort, ID_50_ values correlated with the OD values for IgG and IgM against S and RBD (**Figure S4B**). However, in contrast, the IgM and IgG responses to N in HCW correlated poorly (r^2^ = 0.030 and 0.381 respectively) (**Figure S4B**). Comparison of the peak ID_50_ between asymptomatic individuals, and symptomatic HCWs showed a very similar mean peak ID_50_. In contrast, both groups had lower mean ID_50_ values compared to hospitalized individuals in the 0-3 and 4/5 severity groups (**Figure 5B**). Importantly, some asymptomatic individuals could generate neutralization titres >1,000. Similar to the cohort with confirmed SARS-CoV-2 infection, a decline in ID_50_ was observed following peak neutralization. For many individuals with a peak ID_50_ in the 100-500 range, neutralization was approaching baseline after 50 days POS (**Figure 5C**). As the mean peak ID_50_ was lower in the HCW cohort, the decline in nAb titres towards baseline was more frequent compared to the patient cohort.

**Figure 5:**
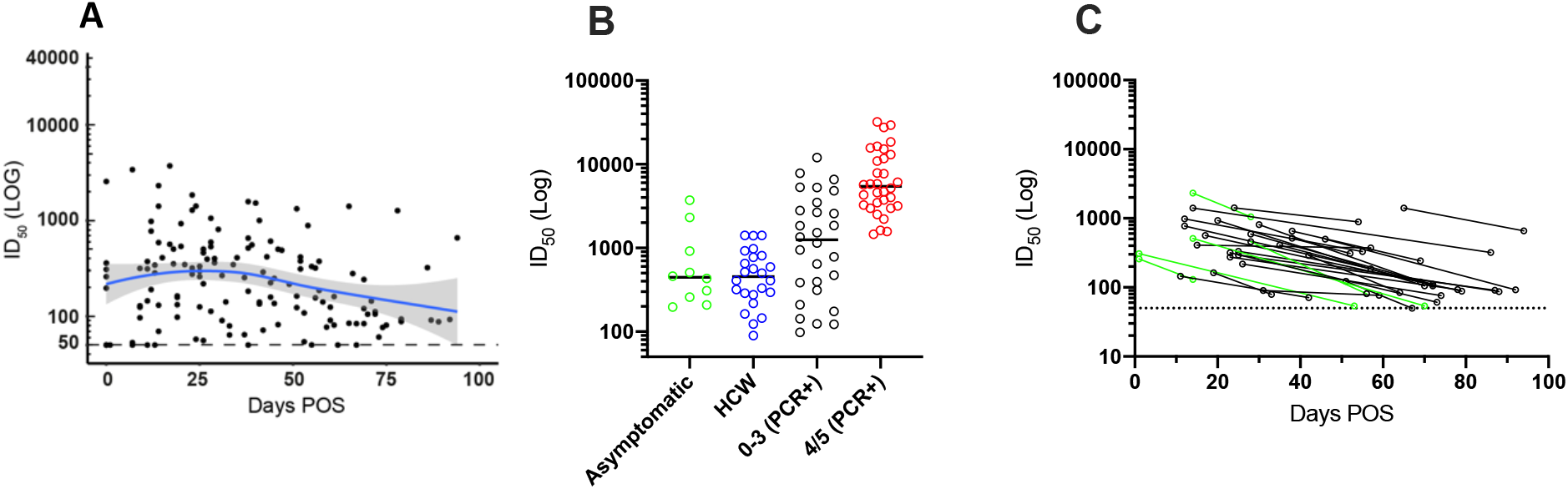
Ab responses in a healthcare worker cohort. A) ID_50_ values plotted against the time post onset of symptoms (POS) at which sera was collected. The line shows the mean ID_50_ value expected from a Loess regression model, the ribbon indicates the pointwise 95% confidence interval. B) Comparison of the peak ID_50_ between asymptomatic individuals (includes 7 HCW and 3 hospital patients), healthcare workers (24 symptomatic HCW with no PCR test), and PCR+ individuals with either severity 0-3 (n=28) or 4/5 (n =32). The 2 PCR+ individuals sampled at early time points (<8 days POS) and did not seroconvert were not included in this analysis. C) ID_50_ at peak neutralization is plotted with the donor matched ID_50_ at the last time point sera was collected. The dotted line represents the cut-off for the pseudotype neutralization assay. Asymptomatic donors are shown in green.

## Discussion

Here, we describe the Ab responses in sequential samples from multiple individuals following SARS-CoV-2 infection in hospitalized patients and healthcare workers. We show that all PCR+ patients sampled >8 days POS developed nAbs with peak ID_50_ in the range of 98-32,000. This wide range in nAb titres against SARS-CoV-2 pseudotyped virus has been observed in other cross-sectional cohorts.^4,16^ Although the average nAb titre was higher in those with more severe disease, the average time to reach peak neutralization did not differ between the 0-3 and 4/5 severity groups. This suggests that disease severity enhances the magnitude of the nAb response but to a lesser extent the kinetics of the nAb response. Importantly, some seropositive individuals who were asymptomatic were able to generate nAb titres >1000. Indeed, highly potent neutralizing monoclonal antibodies (mAbs) have been isolated from asymptomatic patients.^20^ It is not clear why nAb responses correlate with disease severity. A higher viral load may lead to more severe disease and generate a stronger Ab response through increased levels of viral antigen. Alternatively, Abs could have a causative role in disease severity, although there is currently no evidence for antibody dependent enhancement in COVID-19.^21^

Cross-sectional studies in SARS-CoV-2 infected individuals have shown lower mean ID_50_ for serum samples collected at later time points POS (23-52 days).^7^ Longitudinal Abs studies using sequential samples have mostly been limited to 30 days POS.^16^ In two separate studies, IgG binding to S was maintained up until 20-25 days^3^ and day 30 POS^5^. However, a decline in nAb titres have been reported in a small subset of individuals followed sequentially for up to 43 days^22^. The sequential serum samples studied here allowed the measurement of Ab responses up to 94 days POS enabling us to look further into the longevity of the nAb response to SARS-CoV-2 infection in much greater detail than has hitherto been possible. A comparison of the peak ID_50_ value for each individual (mean 23.1 days POS) and ID_50_ at their final timepoint collected, showed a decline in neutralizing titres in both cohorts, regardless of disease severity. This decrease was mirrored in the reduction in IgG binding titres (EC_50_) to S and RBD for the PCR+ cohort (**Figure 4B**). For some individuals with a peak ID_50_ in the 100-300 range, neutralizing titres were at, or below, the level of detection in the SARS-CoV-2 pseudotype neutralization assay after only ∼50 days from the measured peak of neutralization. This trend was also seen in the HCW cohort, and reveals that in some individuals, SARS-CoV-2 infection generates only a transient Ab response that rapidly declines. For those with peak ID_50_ titres >2,000, decline in nAb titres ranged from 2- to 23-fold over an 18-65 day period. It is not clear whether this decline will continue on a downward trajectory or whether the IgG level will plateau to a steady state. Although some nAb titres remain in the 1000-3500 range at the final time point (ranging from 50-82 days POS), further follow up in these cohorts is required to fully assess the longevity of the nAb response in these individuals. Importantly, class-switched IgG memory B cells against S and RBD have been detected in blood of COVID-19 patients showing memory responses are generated during infection.^8,23,24^

The rapid decline observed in IgM and IgA specific responses to S, RBD and N after 20-30 days demonstrates the value of measuring longer lasting SARS-CoV-2 specific IgG in diagnostic tests and seroprevalence studies. However, the waning IgG response should be considered when conducting seroprevalence studies of individuals of unconfirmed PCR+ diagnosed infection or in diagnosis of COVID-19 related syndromes such as PIMS-TS (inflammatory multisystem syndrome temporally associated with SARS-CoV-2).^25^ IgA and IgM could be used as a marker of recent or acute SARS-CoV-2 infection and therefore may be more relevant in a hospital setting. Although a strong correlation between ID_50_ was observed between IgG, IgM and IgA responses against S and RBD, there were still examples where high binding to S and RBD was observed with very little neutralization and therefore care should be taken when using ELISA (or other methods of detecting binding Abs) as a surrogate measurement for neutralization.^26^

The longevity of Ab responses to other coronaviruses have been studied previously.^10-13^ The Ab response following SARS-CoV infection in a cohort of hospitalized patients was shown to peak around day 30^12^ (average titre 1:590) and a general waning of the binding IgG and nAb followed during the 3-year follow up. Low nAb titres of 1:10 were detected in 17/18 individuals after 540 days.^12^ In a second study, low nAb titres (mean titre, 1:28) could still be detected up to 36 months post infection in 89% of individuals.^15^ In contrast to SARS-CoV-2 infection, SARS-CoV infection typically caused more severe disease and asymptomatic, low severity disease were less common. Therefore, the difference in the longevity of the nAb response observed here between SARS-CoV and SARS-CoV-2 infection may relate to the different clinical manifestation of disease between the two viruses.^27^ The more transient Ab responses in the lower disease severity cases in our cohorts reflect more the immune response to endemic seasonal coronaviruses (i.e. those associated with the common cold) which have also been reported to be more transient.^2^ For example, a recent report of 10 individuals studied over a 35-year period showed re-infections with endemic coronaviruses were frequent 12 months after an initial infection.^14^ Further, individuals experimentally infected with endemic alphacoronavirus 229E, generated high Ab titres after 2 weeks but these rapidly declined in the following 11 weeks and by 1 year, the mean Ab titres had reduced further but they were still higher than before the first virus challenge.^10^ Subsequent virus challenge lead to reinfection (as determined by virus shedding) yet individuals showed no cold symptoms.^10^

The nAb titre required for protection from re-infection in humans is not yet understood. Neutralizing monoclonal antibodies (mAbs) isolated from SARS-CoV-2 infected individuals can protect from disease in animal challenge models in a dose dependant manner.^9,28,29^ SARS-CoV-2 infected rhesus macaques, who developed nAbs titres of ∼100 (range 83-197), did not show any clinical signs of illness when challenged 35 days after the first infection.^30^ However, virus was still detected in nasal swabs, albeit 5-logs lower than in primary infection, suggesting immunologic control rather and sterilizing immunity. Similarly, a second study showed rhesus macaques with nAb titres between 8-20 had no clinical signs of disease or detectable virus following re-challenge 28 days after primary infection.^31^ Therefore, although nAb titres are declining over a 2-3 month period in the two cohorts described here, individuals with high peak ID_50_s (>2,000) would likely have sufficient nAb titres to be protected from clinical illness for some time if re-exposed to SARS-CoV-2.

Even though the role of nAbs in viral clearance in primary SARS-CoV-2 infection is not fully understood, many current vaccine design efforts focus on eliciting a robust nAb response to provide protection from infection. Vaccine challenge studies in macaques can give limited insight into nAb titres required for protection from re-infection. Vaccine candidates tested thus far in challenge studies have elicited modest nAb responses (ID_50_ 5-250).^32-35^ For example, a DNA vaccine encoding SARS-CoV-2 S generated nAb titres between 100-200 which were accompanied by a lowering of the viral load by 3-logs. nAb titres in vaccinated animals were shown to strongly correlate with viral load.^34^ However, the role T-cell responses generated through either infection^36^ or vaccination play in controlling disease cannot be discounted in these studies and defining further the correlates and longevity of vaccine protection is needed. Taken together, despite the waning nAb titres in individuals, it is possible that nAb titres will still be sufficient to provide protection from COVID-19 disease for a period of time. However, sequential PCR testing and serology studies in individuals known to have been SARS-CoV-2 infected will be critical for understanding the ability of nAbs to protect from renewed infection in humans.

In summary, using sequential samples from SARS-CoV-2 infected individuals collected up to 94 days POS, we demonstrate declining nAb titres in the majority of individuals. For those with a low nAb response, titres can return to base line over a relatively short period. Further studies using sequential samples from these individuals is required to fully determine the longevity of the nAb response and studies determining the nAb threshold for protection from re-infection are needed.

## Methods

### Ethics

Surplus serum from patient biochemistry samples taken as part of routine care were retrieved at point of discard, aliquoted, stored and linked with a limited clinical dataset by the direct care team, before anonymization. Work was undertaken in accordance with the UK Policy Framework for Health and Social Care Research and approved by the Risk and Assurance Committee at Guy’s and St Thomas’ NHS Foundation Trust (GSTFT). Serum was collected from consenting healthcare workers with expedited approval from GSTFT Research & Development office, Occupational Health department and Medical director.

### Patient and sample origin

269 individual venous serum samples collected at St Thomas’ Hospital, London from 59 patients diagnosed as SARS-CoV-2 positive via real-time RT-PCR, were obtained for serological analysis. Samples ranged from 1 to 94 days after onset of self-reported symptoms or, in asymptomatic cases, days after positive PCR result. Patient information is given in Table S1.

### Healthcare worker (HCW) cohort

Sequential serum samples were collected every 1-2 weeks from healthcare workers at GSTFT between 13^th^ March and 10^th^ June 2020. Seropositivity to SARS-CoV-2 was determined using sera collected in April and early May 2020 using ELISA. Individuals were considered seropositive if sera (diluted 1:50) gave an OD for IgG against both N and S that was 4-fold above the negative control sera.^6^ Self-reported COVID-19 related symptoms were recorded by participants and days post onset of symptoms in seropositive individuals was determined using this information. For asymptomatic, seropositive individuals, days POS was defined as the first timepoint SARS-CoV-2 Abs were detected. Six participants had confirmed PCR+ infection and were included with the PCR+ hospitalized patients in the initial analysis.

### COVID-19 severity classification

Patients diagnosed with COVID-19 were classified as follows:

0 - asymptomatic OR no requirement for supplemental oxygen.

1 - requirement for supplemental oxygen (FiO_2_ <0.4) for at least 12 hrs.

2 - requirement for supplemental oxygen (FiO_2_ ≥0.4) for at least 12 hrs.

3 - requirement for non-invasive ventilation (NIV)/ continuous positive airways pressure (CPAP) OR proning OR supplemental oxygen (FiO_2_ >0.6) for at least 12 hrs AND not a candidate for escalation above level one (ward-based) care.

4 - requirement for intubation and mechanical ventilation OR supplemental oxygen (FiO_2_>0.8) AND peripheral oxygen saturations <90% (with no history of type 2 respiratory failure (T2RF)) OR <85% (with known T2RF) for at least 12 hrs.

5 - requirement for extracorporeal membrane oxygenation (ECMO).

### Protein expression

N protein was obtained from Leo James and Jakub Luptak at LMB, Cambridge. The N protein used is a truncated construct of the SARS-CoV-2 N protein comprising residues 48-365 (both ordered domains with the native linker) with an N terminal uncleavable hexahistidine tag. N was expressed in *E. Coli* using autoinducing media for 7h at 37°C and purified using immobilised metal affinity chromatography (IMAC), size exclusion and heparin chromatography.

S protein consists of a pre-fusion S ectodomain residues 1-1138 with proline substitutions at amino acid positions 986 and 987, a GGGG substitution at the furin cleavage site (amino acids 682-685) and an N terminal T4 trimerisation domain followed by a Strep-tag II.^8^ The plasmid was obtained from Philip Brouwer, Marit van Gils and Rogier Sanders at The University of Amsterdam. The protein was expressed in 1 L HEK-293F cells (Invitrogen) grown in suspension at a density of 1.5 million cells/mL. The culture was transfected with 325 µg of DNA using PEI-Max (1 mg/mL, Polysciences) at a 1:3 ratio. Supernatant was harvested after 7 days and purified using StrepTactinXT Superflow high capacity 50% suspension according to the manufacturer’s protocol by gravity flow (IBA Life Sciences).

The RBD plasmid was obtained from Florian Krammer at Mount Sinai University.^1^ Here the natural N-terminal signal peptide of S is fused to the RBD sequence (319 to 541) and joined to a C-terminal hexahistidine tag. This protein was expressed in 500 mL HEK-293F cells (Invitrogen) at a density of 1.5 million cells/mL. The culture was transfected with 1000 µg of DNA using PEI-Max (1 mg/mL, Polysciences) at a 1:3 ratio. Supernatant was harvested after 7 days and purified using Ni-NTA agarose beads.

### ELISA protocol

ELISA was carried out as previously described.^6^ All sera/plasma were heat-inactivated at 56°C for 30 mins before use in the in-house ELISA. High-binding ELISA plates (Corning, 3690) were coated with antigen (N, S or RBD) at 3 µg/mL (25 µL per well) in PBS, either overnight at 4°C or 2 hr at 37°C. Wells were washed with PBS-T (PBS with 0.05% Tween-20) and then blocked with 100 µL 5% milk in PBS-T for 1 hr at room temperature. Wells were emptied and sera diluted at 1:50 in milk was added and incubated for 2 hr at room temperature. Control reagents included CR3009 (2 µg/mL), CR3022 (0.2 µg/mL), negative control plasma (1:25 dilution), positive control plasma (1:50) and blank wells. Wells were washed with PBS-T. Secondary antibody was added and incubated for 1 hr at room temperature. IgM was detected using Goat-anti-human-IgM-HRP (1:1,000) (Sigma: A6907), IgG was detected using Goat-anti-human-Fc-AP (1:1,000) (Jackson: 109-055-043-JIR) and IgA was detected Goat-anti-human-IgA-HRP (1:1,000) (Sigma: A0295). Wells were washed with PBS-T and either AP substrate (Sigma) was added and read at 405 nm (AP) or 1-step TMB substrate (Thermo Scientific) was added and quenched with 0.5 M H_2_S0_4_ before reading at 450 nm (HRP).

EC_50_ values were measured using a titration of serum starting at 1:50 and using a 5-fold dilution series. Half-maximal binding (EC_50_) was calculated using GraphPad Prism.

### Virus preparation

Pseudotyped HIV virus incorporating the SARS-Cov2 spike protein was produced in a 10 cm dish seeded the day prior with 3.5×10^6^ HEK293T/17 cells in 10 ml of complete Dulbecco’s Modified Eagle’s Medium (DMEM-C) containing 10% (vol/vol) foetal bovine serum (FBS), 100 IU/ml penicillin and 100 μg/ml streptomycin. Cells were transfected using 35 μg of PEI-Max (1 mg/mL, Polysciences) with: 1500 ng of HIV-luciferase plasmid, 1000 ng of HIV 8.91 gag/pol plasmid and 900 ng of SARS-2 spike protein plasmid.^17,18^ The media was changed 18 hours post-transfection and supernatant was harvested 48 hours post-transfection. Pseudotype virus was filtered through a 0.45μm filter and stored at −80°C until required.

### Neutralization assays

Serial dilutions of serum samples (heat inactivated at 56°C for 30mins) were prepared with DMEM media and incubated with pseudotype virus for 1-hour at 37°C in 96-well plates. Next, Hela cells stably expressing the ACE2 receptor (provided by Dr James Voss, The Scripps Research Institute) were added and the plates were left for 72 hours. Infection level was assessed in lysed cells with the Bright-Glo luciferase kit (Promega), using a Victor™ X3 multilabel reader (Perkin Elmer).

### Statistical analysis

Analyses were performed using R (version 4.0.0) and GraphPad Prism (version 7.0.4). On charts showing OD/ID_50_ and days post-infection, the overall trend in the data was indicated by lines generated using Loess regressions (span 1.5) with ribbons depicting the 95% confidence intervals.

**Table 1:** Cohort description. Gender, severity, age, and outcome.

| **Gender** |  |
| --- | --- |
| Male | 51 (78.5%) |
| Female | 14 (21.5%) |
| **Age** |  |
| Mean | 55.2 years (23-95) |
| **Severity** |  |
| 0 | 14 |
| 1 | 10 |
| 2 | 7 |
| 3 | 2 |
| 4 | 25 |
| 5 | 7 |
| **Outcome** |  |
| HCW | 6 |
| Died | 12 |
| Discharged | 41 |
| Still in hospital | 5 |
| Transferred to local hospital | 3 |

## Data Availability

Anonymised data was used in this study and is not available for distribution outside the host organisations.

## Acknowledgements

King’s Together Rapid COVID-19 Call awards to MHM, KJD, SJDN and RMN.

MRC Discovery Award MC/PC/15068 to SJDN, KJD and MHM.

AWS and CG were supported by the MRC-KCL Doctoral Training Partnership in Biomedical Sciences (MR/N013700/1).

GB was supported by the Wellcome Trust (106223/Z/14/Z to MHM).

SA was supported by an MRC-KCL Doctoral Training Partnership in Biomedical Sciences industrial Collaborative Award in Science & Engineering (iCASE) in partnership with Orchard Therapeutics (MR/R015643/1).

NK was supported by the Medical Research Council (MR/S023747/1 to MHM).

SP, HDW and SJDN were supported by a Wellcome Trust Senior Fellowship (WT098049AIA). Fondation Dormeur, Vaduz for funding equipment (to KJD).

Development of SARS-CoV-2 reagents (RBD) was partially supported by the NIAID Centers of Excellence for Influenza Research and Surveillance (CEIRS) contract HHSN272201400008C.

Thank you to Florian Krammer for provision of the RBD expression plasmid, Philip Brouwer, Marit Van Gils and Rogier Sanders (University of Amsterdam) for the S protein construct, Leo James, Jakub Luptak and Leo Kiss (LMB) for the provision of purified N protein, and James Voss for providing the Hela-ACE2 cells. We that Isabella Huettner for assistance with figures. We are extremely grateful to all patients and staff at St Thomas’ Hospital who participated in this study.

We thank the COVID-19 core research team members including Olawale Tijani, Kate Brooks, Michael Flanagan, Robert Kaye, Raenelle Williams, Cristina Blanco-Gil, Helen Kerslake, Annelle Walters, Rizwana Dakari, Jennifer Squires, Anna Stanton, Sherill Tripoli, Andrew Amon, Isabelle Chow and Olanike Okolo.

